# A Simplified Point-of-Care Lung Ultrasound Protocol to Detect Coronavirus Disease 2019 in Inpatients: A Prospective Observational Study

**DOI:** 10.1101/2021.04.19.21254974

**Authors:** Thomas F. Heyne, Benjamin P. Geisler, Kay Negishi, Daniel Choi, Ahad A. Al Saud, Lucas X. Marinacci, Patrick Y. Smithedajkul, Lily R. Devaraj, Brent P. Little, Dexter P. Mendoza, Efren J. Flores, Milena Petranovic, Steven P. Toal, Hamid Shokoohi, Andrew S. Liteplo

## Abstract

**Objectives:** To assess the diagnostic performance of lung point-of-care ultrasound (POCUS) compared to either a positive nucleic acid test (NAT) or a COVID-19-typical pattern on computed tomography (CT) and to evaluate opportunities to simplify a POCUS algorithm.

**Methods:** Hospital-admitted adult inpatients with (1) either confirmed or suspected COVID-19 and (2) a completed or ordered CT within the preceding 24 hours were recruited. Twelve lung zones were scanned with a handheld POCUS machine. POCUS, CT, and X-ray (CXR) images were reviewed independently by blinded experts. A simplified POCUS algorithm was developed via machine learning.

**Results:** Of 79 enrolled subjects, 26.6% had a positive NAT and 31.6% had a CT typical for COVID-19. The receiver operator curve (ROC) for a 12-zone POCUS protocol had an area under the curve (AUC) of 0.787 for positive NAT and 0.820 for typical CT. A simplified four-zone protocol had an AUC of 0.862 for typical CT and 0.862 for positive NAT. CT had an AUC of 0.815 for positive NAT; CXR had AUCs of 0.793 for positive NAT and 0.733 for typical CT. Performance of the four-zone protocol was superior to CXR for positive NAT (p=0.0471). Using a two-point cutoff system, the four-zone POCUS protocol had a sensitivity of 0.920 and 0.904 compared to CT and NAT, respectively, at the lower cutoff; it had a specificity of 0.926 and 0.948 at the higher cutoff, respectively.

**Conclusion:** POCUS outperformed CXR to predict positive NAT. POCUS could potentially replace other chest imaging for persons under investigation for COVID-19.

## INTRODUCTION

Rapid and accurate diagnosis of Coronavirus Disease 2019 (COVID-19) can be challenging. The most widely used diagnostic test to detect Severe Acute Respiratory Syndrome Coronavirus 2 (SARS-CoV-2) infection is a nucleic acid amplification test (NAT) via reverse-transcription polymerase chain reaction. NAT via nasopharyngeal swab has a very high specificity but a sensitivity of only 70-85%.^1^ Thus, a significant number of patients may be infected even with a negative NAT, especially later in the course of infection.^2,3^ Computed tomography (CT) of the chest, which can detect typical patterns of lung findings for COVID-19, has a higher sensitivity (86-97%) but lower specificity (25-81%) compared to NAT.^4–7^ Several validated reporting systems are used by radiologists in assessing likelihood of COVID-19 pneumonia on chest CT. A commonly used consensus guideline from the Radiological Society of North America (RSNA) classifies CT morphologies as “typical” (highest suspicion), “indeterminate”, “atypical”, or “negative” for COVID-19 pneumonia.^8^ However, CT scan has drawbacks; it requires significant healthcare resources and time, exposes the patient to ionizing radiation, poses infection control risks, and may be unsafe for unstable, hypoxemic patients. Chest X-ray (CXR) reduces some of these drawbacks but has a low sensitivity for detecting COVID-19, particularly early in the disease course (55-83%).^9^ Thus, hospitals often rely on more than one category of diagnostic test, as well as patient history and risk factors, to attempt to rule out COVID-19 infection. Imaging, therefore, can be a helpful adjunct.^6,10^

In our hospital system, either a CXR or a CT is required for all admitted patients with a clinical suspicion for COVID-19 (based on history and demographics) and a negative swab.^11^ If CXR findings are concerning for COVID-19, a CT scan and second NAT are often recommended. If the CT scan does not show a “typical” pattern for COVID-19, and the second NAT is negative, then COVID-19 is usually ruled out. While the case remains uncertain (while patients are still undergoing diagnostic evaluation for suspected COVID-19), patients are flagged as Persons Under Investigation (PUIs) and placed under the same enhanced isolation precautions as patients with COVID-19. Thus, both CXR and CT scan can help in ruling out and ruling in COVID-19, but they have drawbacks.

Point-of-care ultrasound (POCUS) may be an alternative diagnostic modality. POCUS is inexpensive, rapid, safe (no radiation), widely available, and does not require travel to and possible infectious exposure of a radiology suite. COVID-19 tends to affect the lung periphery, which is visualized well by POCUS. Numerous studies have described characteristic features for COVID-19 on lung ultrasound, including confluent B-lines (also known as “waterfall” B-line, “light beam artifact”, or “white” lung).^12–15^ Multiple stratifying scoring systems to assess disease severity have been proposed. For example, an expert opinion from Soldati et al. proposed scanning 14 lung zones and assigning 0-3 points per zone; the total number of points would correlate with the severity of COVID-19 pneumonia.^16^ The same group analyzed protocols with fewer lung zones (e.g., 4-12 zones), given that an abbreviated protocol could help reduce exposure time with infectious patients; however, the study concluded that a 12-zone protocol was optimal to assess COVID-19 severity.^17^ To date, relatively few prospective research studies have evaluated the accuracy of POCUS to assist in the diagnosis of COVID-19.^18–20^ While valuable, these studies have limitations which hinder their generalizability, including using a single operator, excluding patients with heart failure, or relying on the scanner’s overall subjective gestalt for COVID-19 pneumonia.

Our aim was threefold. First, we assessed the diagnostic accuracy of lung POCUS to detect either (1) a high-suspicion (“typical”) pattern for COVID-19 on CT scan or (2) a positive COVID-19 NAT. We hypothesized that lung POCUS has superior accuracy compared to CXR for both outcomes. Second, we assessed whether diagnostic accuracy would be affected by a simplified scanning protocol, which required fewer lung zones and less subtle findings than the protocol proposed by Soldati et al.^16^ Third, we assessed whether diagnostic accuracy would differ in the subpopulation of patients with a diagnosis of congestive heart failure (CHF), given possible overlap in findings between COVID-19 and CHF, and therefore possible reduced accuracy in this population.

## MATERIALS AND METHODS

### Study Setting and Patient Population

This prospective cohort study took place in a 1,000-bed quaternary care hospital, from April to July 2020. We included a convenience sample of adult patients who were admitted to either the medical ward or intensive care unit (ICU). A query of the electronic medical record system identified patients who had either confirmed or suspected COVID-19 infection (i.e., “PUI” status and awaiting further testing) and who had completed or were planned to complete a chest CT within 24 hours of the POCUS scan. Patients with interstitial lung disease (ILD) were excluded, given expected abnormal lung POCUS scans. The protocol was approved in writing by the local Institutional Review Board.

### Data Collection

After obtaining assent from the treating team and verbal consent from the patient or proxy, a research physician scanned 12 lung zones (Figure 1), similar to prior protocols.^20^ The patient wore a surgical mask, and the scanning physician wore hospital-recommended personal protective equipment (N-95 respirator, eye protection, gown, and gloves). The probe was held longitudinally and perpendicular to the ribs to obtain the “bat sign” view.^21^ Scanning physicians attempted to capture at least two intercostal spaces for each zone (and at least three intercostal spaces for each of the four posterior zones). For posterior lung zones, patients either sat upright or lay in lateral decubitus position. Scanning was completed using a handheld Butterfly iQ ultrasound machine (Butterfly Network, Inc., Guilford, CT) connected to an iPhone (Apple Inc., Cupertino, CA). The lung preset was used for all zones; a second clip was obtained using the abdominal preset for zones R4 and L4 (to investigate for pleural effusion). Single-use gel packets were used for ultrasound gel, to avoid cross-contamination. After scanning, machines were sanitized with hospital-approved disinfectant wipes. Scanning physicians included four internal medicine and two emergency medicine physicians, all of whom had completed formal POCUS training, including a minimum of 25 lung scans reviewed by a POCUS expert. All scanners were blinded to CT results, though not to NAT results (positive results are prominently displayed in the chart), at the time of scanning. Patient demographics, vital signs, amount of supplemental oxygen, and lab values were recorded by non-scanning research staff.

**Figure 1:**
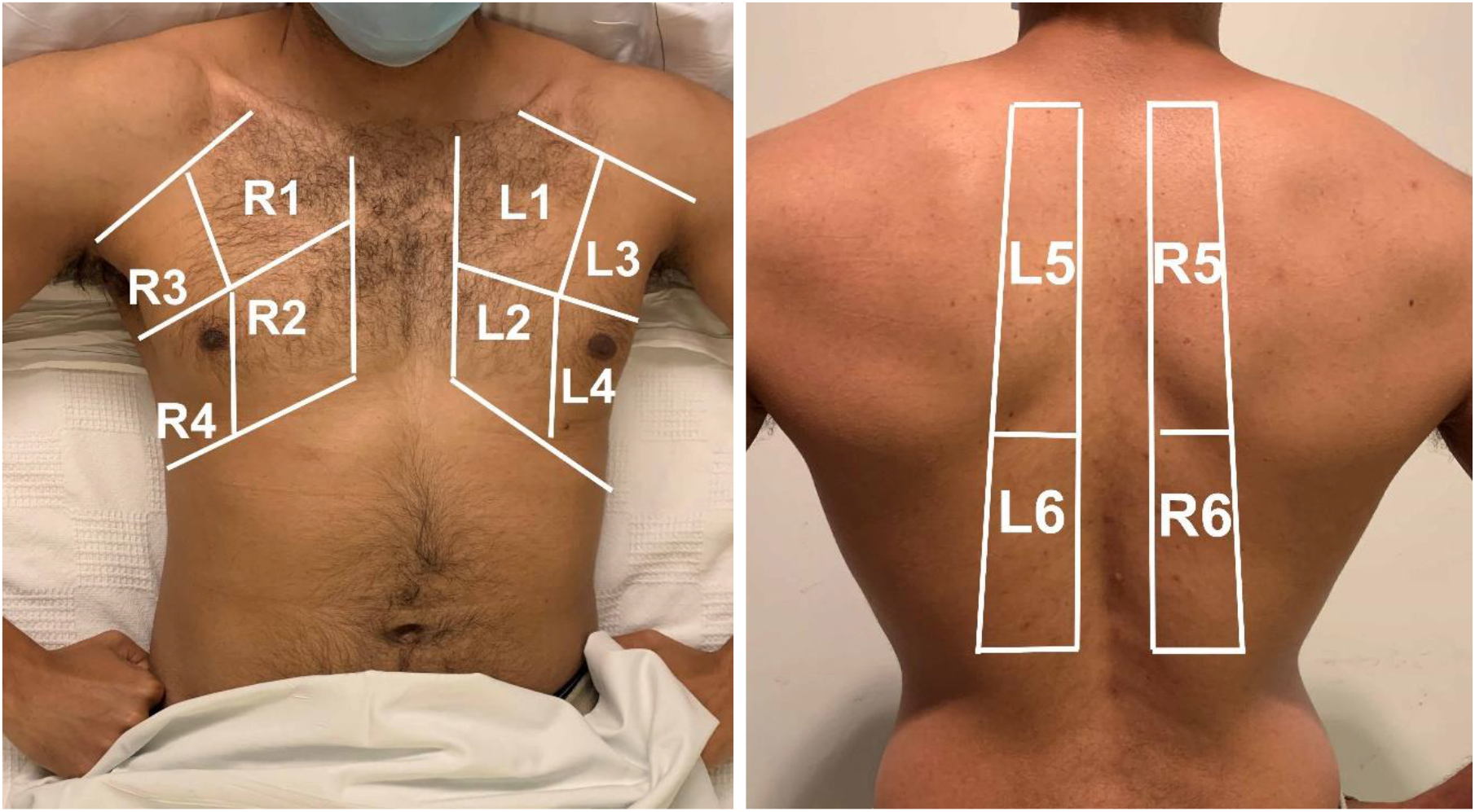
Lung Ultrasound Zones. Anterior lung zones (R1, R2, L1, L2) were scanned along the mid-clavicular line; lateral zones (R3, R3, L3, L4) were scanned along the mid-axillary line; the fourth rib separated the upper and lower anterior-lateral zones. Posterior lung zones (R5, R6, L5, L6) were scanned between the spine and the scapula; upper and lower posterior zones were separated by the inferior border of the scapula. For our simplified protocol, only the four posterior zones are scanned.

POCUS clips were reviewed by two fellowship-trained POCUS experts, who were blinded to CT and NAT results. Scans were scored along numerous criteria, including categories of B-lines, pleural line irregularity, and consolidation, selecting the most severe pathology in each of these 3 categories for each lung zone (see Appendix). B-line examples are pictured (Figure 2). Similarly, CXR and CT scans were reviewed by two board-certified radiologists specialized in thoracic imaging and blinded to clinical and POCUS information. CXRs and CTs were interpreted independently. Radiologists gave each CT and CXR a COVID-19 suspicion grade, following RSNA consensus categories: (1) typical (highest suspicion), (2) indeterminate, (3) atypical/low, or (4) negative.^22^ A third radiologist provided an interpretation in the case of discordant interpretations (as a tiebreaker). For both CXR and CT, the mode of the three interpretations was considered the consensus. For cases in which the third reader assigned a grade different from the primary two readers, the grade corresponding to the value closest to the median of the three observer grades was used as the consensus.

**Figure 2:**
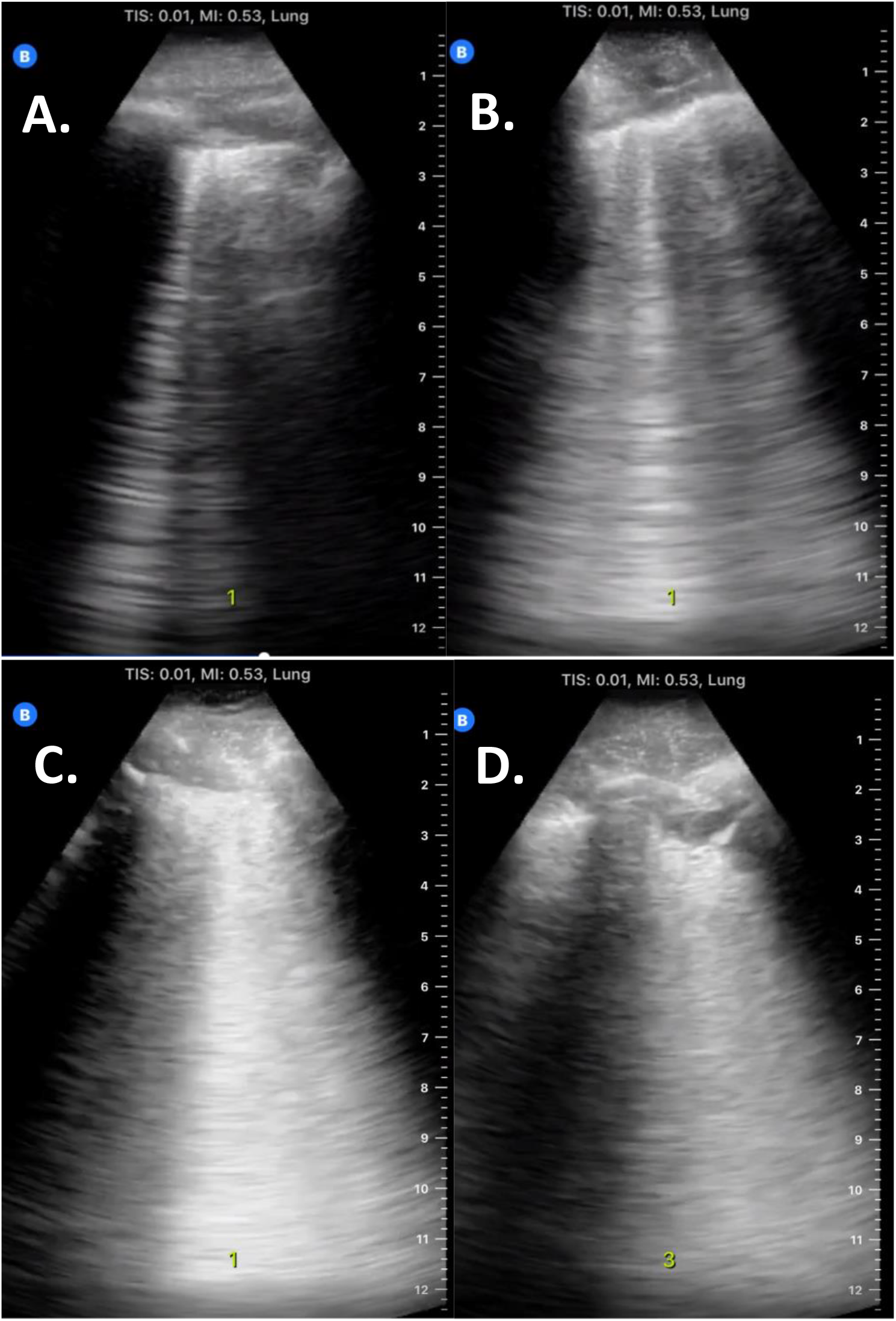
Examples of B-lines. **A**. 1-2 B-lines. **B**. 3+ B-lines in one intercostal space. **C**. Confluent or coalescing B-lines. **D**. Comet tail artifacts coming below a subpleural consolidation (arising significantly below pleural line); these were <u>not</u> counted as B-lines. B-lines were only scored if arising from/close to the pleural line (approximately ≤ 4 mm from the pleural line).

### Data Analysis

For all imaging modalities, we calculated a Cohen’s kappa to compare the interrater reliability between the first two readers (not including the “tiebreaker”). An ordinal scale was used for CT, CXR, and POCUS interpretations. For POCUS scans, we recorded the duration of time spent scanning (from the start of the first clip to the final clip).

#### Accuracy of 12-Zone Soldati Protocol

We assessed the accuracy of the 12-zone Soldati POCUS protocol to detect either (1) typical CT pattern or (2) positive NAT (by the time of discharge). The Soldati protocol assigned a score 0-3 for each of the 12 lung zones (thus, maximum score for a patient was 36).^16^ Receiver operating characteristic (ROC) curves were created to compare the performance of Soldati score and CXR for our first outcome: a typical pattern for COVID-19 on CT. Additional ROC curves compared Soldati score, CXR, and CT against our second outcome: positive NAT. Areas under the curve (AUCs) were generated with the algorithm suggested by DeLong, DeLong, and Clark-Pearson.^23^

#### Creation and Accuracy of the Simplified POCUS Protocol

We next evaluated simplified modifications of the Soldati protocol. Machine learning analyses employing random forests and lasso regression were used to identify the lung zones and findings most correlative to our two outcomes (typical CT pattern and positive NAT). Using this information, we generated modified (simplified) versions of the Soldati protocol and analyzed the accuracy of each modification. Specifically, we sequentially reduced the number of lung zones that were incorporated into the POCUS score (starting at 12 and ending at 4 zones). In addition, we modified the point values that the Soldati score had assigned to different lung findings. For example, we varied whether 0-4 points would be scored for each lung zone that showed 3+ B-lines. In total, over 150 scoring algorithms (modifications of the original Soldati score) were generated. ROC curves were generated for each of these modified scoring systems for the outcomes of (1) typical CT pattern and (2) positive NAT. The AUCs for each of these modified scores were compared to both the Soldati Score and the other imaging modalities (CXR for typical CT pattern, and CXR and CT for positive NAT). Finally, we performed the same analysis (ROC curves for our two outcomes) in the subpopulation of patients with a history of CHF.

To calculate individual test characteristics (sensitivity, specificity, positive and negative predictive value) for our simplified POCUS score, we created 2 score cutoffs. These cutoffs allowed us to create low, intermediate, and high COVID-suspicion categories from the ordinal POCUS score (e.g., 0-12 points); these categories would mirror the RSNA CT and CXR categories.

Data analysis was performed in Python 3.8.3 (Python Software Foundation, Beaverton, OR)^24^ and STATA IC 14.2 (Stata Corp LLC, College Station, TX).

## RESULTS

Ninety patients were initially screened, and 79 were scanned and included in the final analysis. The remaining eleven were excluded based on lack of consent, presence of exclusion criteria (namely ILD), or CT scan not completed (see Appendix). Patient characteristics at the time of scan are given in Table 1. Most subjects were male (67.1%), of advanced age (mean age 62.5 years), overweight (mean BMI 27.0 kg/m^2^), and with elevated inflammatory markers.

**Table 1:** Patient Characteristics at the Time of POCUS Scan.

|  | **All**<br>**n=79** | **COVID-19**<br>**NAT positive**<br>**n=21 (26.6%)** | **COVID-19**<br>**NAT negative**<br>**n=58 (73.4%)** | **p-**<br>**value** |
| --- | --- | --- | --- | --- |
| Age (years);<br>mean (95% CI) | 62.5<br>(58.6-66.4) | 58.6<br>(50.0-67.2) | 63.9<br>(59.7-68.2) | 0.231 |
| Gender female; % | 32.9% | 28.6% | 34.5% | 0.788 |
| Time since symptom onset<br>(days); mean (95% CI) | 8.8<br>(5.5-12.2) | 10.0<br>(8.8-11.3) | 10.1<br>(9.2-10.9) | 0.055 |
| Scanned in ICU; % | 5.1% | 14.3% | 1.7% | 0.055 |
| Body mass index (kg/m<sup>2</sup>);<br>median (95% CI) | 26<br>(25-27.7) | 29<br>(25.5-31) | 25.5<br>(24-26.9) | 0.125 |
| History of CHF; % | 21.5% | 14.3% | 24.1% | 0.537 |
| Supplemental O<sub>2</sub> (L/min)*;<br>mean (95% CI) | 2.7<br>(2.2-3.3) | 2.6<br>(1.2-3.9) | 2.8<br>(2.3-3.4) | 0.670 |
| Absolute lymphocyte count<br>(per μL); median (95% CI) | 1,125<br>(972-1,360) | 1,280<br>(814-1,545) | 1,110<br>(943-1,447) | 0.710 |
| Ferritin (ng/mL);<br>median (95% CI) | 353<br>(230-510) | 691<br>(426-870) | 220<br>(142-353) | 0.001 |
| CRP (mg/L);<br>median (95% CI) | 62.1<br>(38.5-84.8) | 71.8<br>(48.7-132.8) | 52.8<br>(11.6-85.9) | 0.145 |
| LDH (U/L);<br>median (95% CI) | 258<br>(226-326) | 327<br>(266-394) | 224<br>(180-258) | 0.003 |
| D-dimer (ng/mL);<br>median (95% CI) | 1,341<br>(1,066-1,577) | 1,431<br>(891-2,435) | 1,314<br>(1,010-1,573) | 0.738 |
| NT-proBNP (pg/mL);<br>median (95% CI) | 548<br>(307-2,044) | 307<br>(34-1,843) | 600<br>(322-2,844) | 0.139 |
| In-hospital death; % | 7.6% | 14.3% | 5.2% | 0.333 |
| Discharged to hospice; % | 1.3% | 0.0% | 1.7% | 1.000 |

\*Includes only 28 patients on nasal cannula.
Abbreviations: NAT, nucleic acid test; CI; confidence interval; CHF, congestive heart failure; ICU, intensive care unit; CRP, C-reactive protein; LDH, Lactate dehydrogenase; NT pro-BNP, N-terminal pro B-type natriuretic peptide.

Overall, 26.6% (21/79) of patients were NAT positive. All tested positive on their initial NAT. No patient who had an initial negative NAT tested subsequently tested positive during the study period. For CT scan, patients received the following RSNA grades (consensus interpretation): 18.9% (15/79) negative, 27.8% atypical/low (22/79), 21.5% indeterminate (17/79), and 31.6% typical (25/79) (see Appendix). Seventy-five patients (94.9%) had CXRs completed; of these, the radiology consensus was 25.3% (19/75) negative, 24.0% (18/75) atypical/low, 29.3% indeterminate (22/75), and 21.3% (16/75) typical for COVID-19. Interrater reliability between the two readers for each of the imaging modalities was as follows: κ = 0.822 for CT scan, κ = 0.559 for CXR, κ = 0.704 for the 12-zone Soldati Protocol, and κ = 0.740 for the simplified four-zone protocol (below). The median time between POCUS exam and CT scan completion was 13 hours (95% confidence interval [CI]: 11.1, 16.8). For ten patients, >24 hours elapsed between scans, typically from delays in obtaining CT scan after initial order. The POCUS exam took a median of 10 (95% CI: 9.4; 10.8) minutes. There was no statistically significant difference in the POCUS scan time between NAT-positive and negative patients (p=0.845).

### Accuracy of 12-Zone Soldati Score and Simplified 4-Zone Score

ROC curves were generated, and AUCs were calculated, to assess the accuracy of the 12-zone Soldati protocol for detecting (1) typical CT pattern and (2) positive NAT (Figure 3).

**Figure 3:**
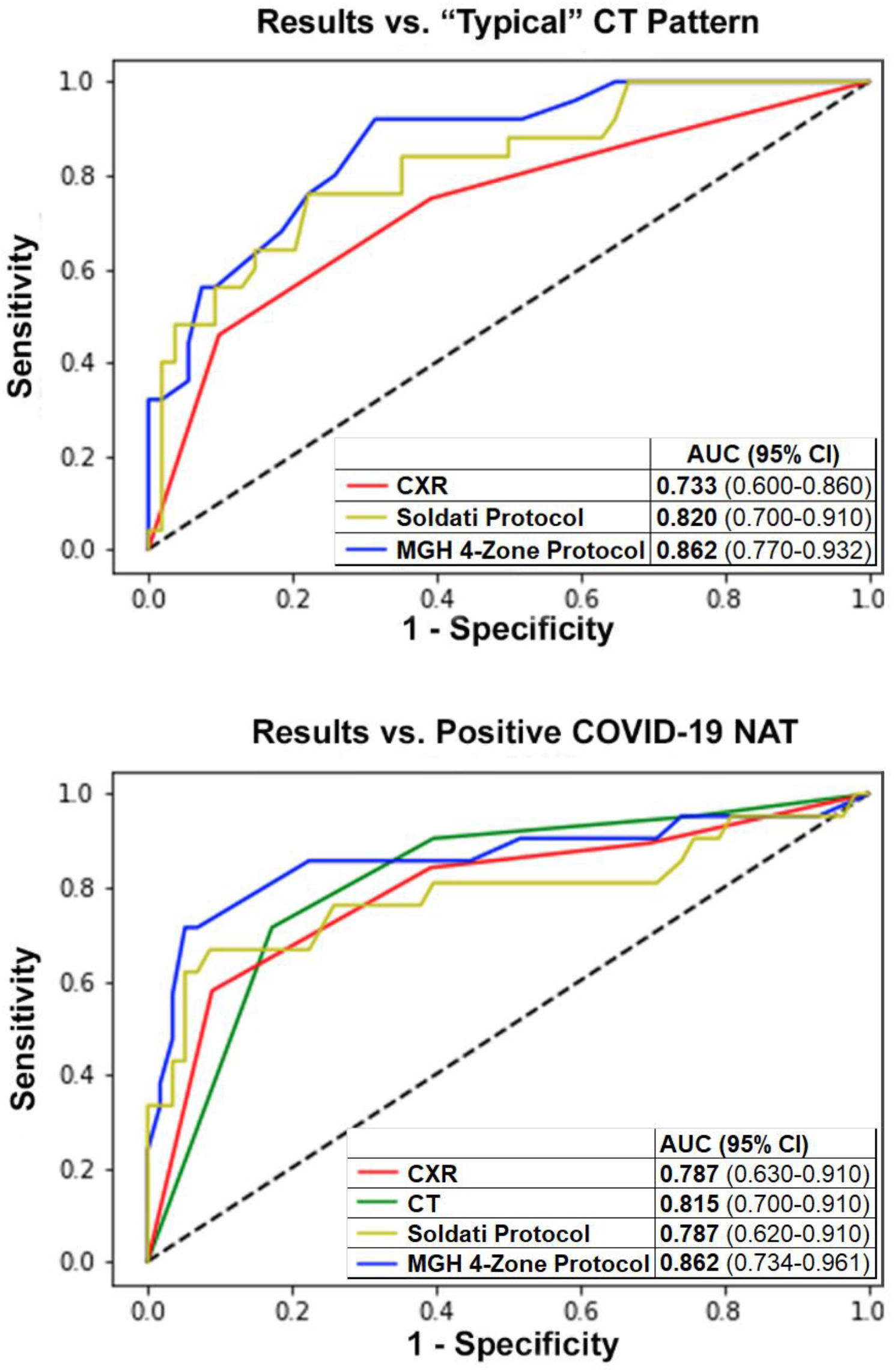
Receiver Operating Characteristic Curves. Abbreviations: CT, computed tomography; CXR, chest X-ray; AUC, area under the curve; CI, confidence interval; NAT, nucleic acid test; MGH, Massachusetts General Hospital

Machine learning analyses identified the four posterior lung zones as having relatively higher feature importance (see Appendix). Among the different lung findings, B-lines of various categories (confluent B-lines, 3+ B-lines, and 1-2 B-lines) had relatively higher feature importance (see Appendix). ROC curves were generated for over 150 variations of the Soldati score, using the methodology described above. For example, one permutation assigned 1 point for 1-2 B-lines, 3 points for confluent B-lines, and 2 points for most other abnormalities (3+ B-lines, pleural line irregularity, any subpleural consolidation, or hepatization). For the outcome of typical CT pattern, this algorithm yielded a ROC with AUC of 0.833 if all 12 zones were included, and AUC of 0.838 if only the four posterior zones were included. However, this algorithm was still relatively complicated.

Our simplest protocol required only four posterior lung zones and assigned 1 point for 1-2 B-lines, 2 points for 3+ B-lines, and 3 points for any confluent B-lines. No points were given for pleural line irregularity or consolidation. We have adopted this four-zone, B-line only protocol as our MGH 4-Zone Protocol (Figure 4). The ROC curves and corresponding AUCs are given (Figure 3). These AUCs were numerically higher than the AUCs for the other diagnostic modalities, but there was only a statistically significant difference between the MGH 4-Zone Protocol and CXR, for the outcome of positive NAT (p = 0.0471). The difference between the MGH 4-Zone Protocol and CXR for the outcome of typical CT pattern reached only borderline statistical significance (p= 0.0930).

**Figure 4:**
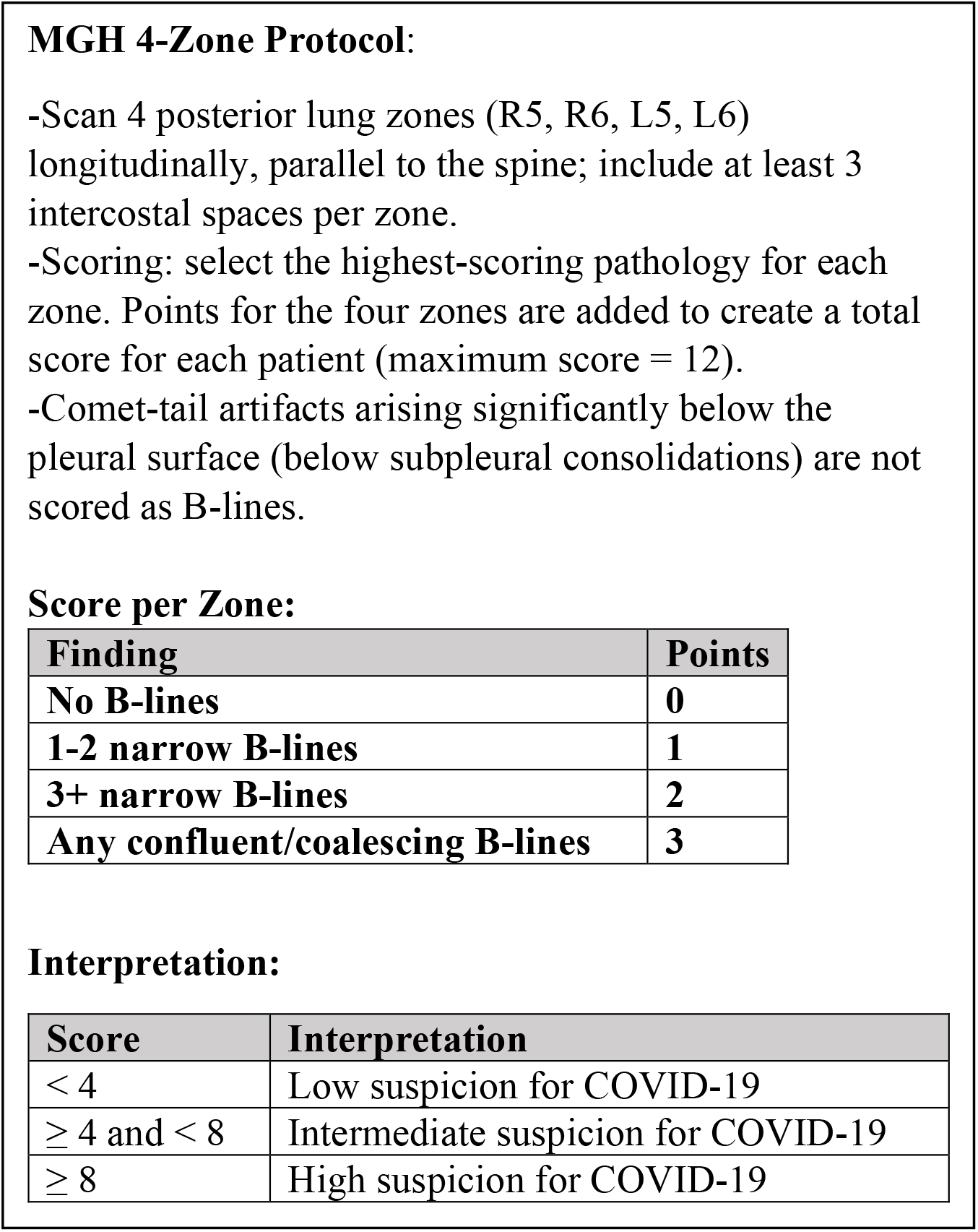
Simplified Four-Zone Protocol.

### Sensitivity, Specificity Using Score Cutoffs

To mirror the RNSA categories used for CT and CXR, we created 2 cutoffs for our MGH 4-Zone Protocol, to divide patients into low, intermediate, and high suspicion for COVID (see Appendix). We set low suspicion at <4 points; intermediate at ≥ 4 and < 8 points, and high suspicion at ≥8 points. Operating characteristics are summarized in Table 2, which also includes the sub-analysis of patients with a prior medical history of CHF (n=17).

**Table 2:** Test Characteristics of MGH 4-Zone Protocol.

|  | **Sensitivity** | **Specificity** | **PPV** | **NPV** |
| --- | --- | --- | --- | --- |
| **All Patients (n=79)** |  |  |  |  |
| For “Typical” CT Pattern: |  |  |  |  |
| Score $\geq 8$ | 0.560 | 0.926 | 0.778 | 0.820 |
| Score $\geq 4$ | 0.920 | 0.537 | 0.479 | 0.935 |
| For Positive NAT: |  |  |  |  |
| Score $\geq 8$ | 0.714 | 0.948 | 0.833 | 0.901 |
| Score $\geq 4$ | 0.905 | 0.500 | 0.396 | 0.935 |
| **Patients with CHF (n=17)** |  |  |  |  |
| For “Typical” CT Pattern: |  |  |  |  |
| Score $\geq 8$ | 0.600 | 0.917 | 0.750 | 0.846 |
| Score $\geq 4$ | 1.000 | 0.583 | 0.500 | 1.000 |
| For Positive NAT: |  |  |  |  |
| Score $\geq 8$ | 1.000 | 0.928 | 0.750 | 1.000 |
| Score $\geq 4$ | 1.000 | 0.500 | 0.300 | 1.000 |

Abbreviations: MGH, Massachusetts General Hospital; CT, computed tomography; NAT, nucleic acid test; CHF, congestive heart failure

## DISCUSSION

A 12-zone lung POCUS protocol following Soldati et al. had reasonable diagnostic accuracy to detect either a typical CT pattern or a positive NAT for COVID-19. This accuracy did not decrease with a simplified four-zone POCUS protocol, which yielded a specificity of more than 90% at the high cut-off point for detecting either typical CT pattern or positive NAT. At the intermediate cut-off point, the sensitivity and negative predictive value for typical CT pattern and for positive NAT were both greater than 90%. When predicting the COVID-19 NAT result, POCUS performed better than chest X-ray. Otherwise, there was no statistically significant difference in the diagnostic accuracy among the 12-zone Soldati protocol, our four-zone protocol, CT, and CXR. We suspect that our simplified four-zone protocol performed well compared to the 12-zone Soldati protocol, because the latter was intended to assess the severity of COVID-19 (e.g., assigning higher points for consolidation), not to diagnose COVID-19. Extrapolating from our results, patients with a high four-zone POCUS score could be considered high risk for COVID-19, and further imaging with a CT could likely be avoided. Indeed, all patients above the high cut-off had either “typical” (14/18) or “indeterminate” (4/18) CT findings. Similarly, one could extrapolate that patients below the low cut-off could be considered to have a low risk for COVID-19 and might be able to forgo a CT scan as well. The protocol performed equally well in CHF patients. In addition, our patients on average were overweight (mean BMI 27.0), suggesting that the protocol would perform well in the overall more obese U.S. population.

The study results add to the literature that POCUS can assist in the diagnosis or risk stratification of PUIs for COVID-19.^18–20^ Namely, lung POCUS may be a helpful adjunct to history and risk factors both to “rule in” or “rule out” COVID-19 infection. POCUS may be particularly helpful in resource-limited settings, where access to CT or NAT may be limited or results may be delayed.

Benefits of lung POCUS include safety for the patient: no need for ionizing radiation (particularly important for younger or pregnant patients) and no need for transportation to a radiology suite (may be unsafe in sicker patients and a potential infectious control risk). One strength of our study is that we used an inexpensive, handheld, easily disinfected ultrasound machine. Such technology is increasingly available worldwide. Also, lung POCUS is rapid. Our 12-zone POCUS mean scanning time was 10 minutes; a protocol with only four zones would have been even faster.

Indeed, strengths of our protocol include simplicity and potential applicability worldwide. The simplified protocol requires scanning only four posterior lung zones using a simple longitudinal technique, meaning scanning less time and standing behind the patient’s face—reducing exposure risk to the scanner. The interpretation is also simpler: it is easier to assess for B-lines than for subtle consolidations or pleural line breaks or irregularities (e.g., as recommended in prior studies).^16,20^ The protocol fits with findings from prior studies, as confluent B-lines (or “light beam artifacts”) have been described as more specific for COVID-19,^12–15^ and as COVID-19 tends to affect the posterior lungs preferentially.^13,17,19,20^ The inter-rater reliability for readers for the simplified POCUS score was overall good, notably higher than CXR and similar to CT.

Our study was subject to several limitations. First, this was a single-center study, using a convenience sample of inpatients based on scanning physician availability. However, there have been relatively few prospective studies to date, particularly multi-center studies. Validation studies performed at other centers are warranted and welcome. Second, our study examined only inpatients (mostly from the ward, with a small number of ICU patients); it is unknown whether this scanning protocol would yield similar results for outpatients or ED patients. Third, the relatively high incidence of COVID-19 (26.6%) could lead to a spectrum effect. However, this incidence is lower than some other studies, and our sample did include many patients with low-suspicion CXR and CT results. Fourth, we note that, although many of our patients were quite sick, none of them were proned at the time of exam. It is unknown whether our protocol would have the same accuracy in proned patients. However, it is rare to prone a ward patient without a confirmed diagnosis of COVID-19. Fifth, although scanning physicians were blinded to CT results, they were not blinded to NAT status (positive COVID status is displayed prominently in our hospital electronic record); it is possible that sampling bias of the lung zones was introduced. However, COVID-positive and PUI patients had similar POCUS scan times. Sixth, none of the patients with an initial negative NAT subsequently had a positive NAT. However, this was an inpatient study, and initial NAT is always performed in the Emergency Department in our institution. Incidence of positive NAT after initial negative NAT is very low.^2^ Thus, it would be extremely challenging to capture an appreciable number of NAT-positive patients if we had limited our study to only those patients with an initial negative NAT. Finally, we acknowledge that many hospitals do not have inpatient providers who are sufficiently trained in lung POCUS. However, interest in POCUS among inpatient clinicians is growing, and applications like these could bring impetus to further clinician training. In addition, most Emergency Department providers in the U.S. are trained in lung POCUS.

In conclusion, lung POCUS could be a valuable tool in assessing patient risk of COVID-19. Both a 12-zone protocol and a simplified four-zone protocol performed well to detect patients with either a high-suspicion CT pattern for COVID-19 or a positive nasopharyngeal NAT. Indeed, the simplified four-zone lung POCUS out-performed CXR for identifying patients with positive NAT. In patients under investigation for COVID-19, a lung POCUS exam could potentially replace other chest imaging as a helpful adjunct for clinicians to decide whether a COVID-19 diagnosis should be further entertained or if patients can be taken off precautions. Further research in the form of confirmatory studies at other centers is warranted.

## Data Availability

The data that support the findings of this study are available on request from the corresponding author, TFH.

## ACKNOWLEDGEMENTS

We greatly appreciate assistance from Dr. Irene Ma and Dr. Emily Hyle for advice in planning this project and revising the manuscript. We are also grateful to Dr. Nicole Duggan and Dr. Daniel Restrepo for assistance in early stages of this project. Preliminary results from this study were presented at the Massachusetts General Hospital Clinical Research Day, a local research meeting held on October 1, 2020.

## REFERENCES

1. Yang Y, Yang M, Shen C, et al. Evaluating the accuracy of different respiratory specimens in the laboratory diagnosis and monitoring the viral shedding of 2019-nCoV infections. medRxiv 2020:2020.02.11.20021493. https://doi.org10.1101/2020.02.11.20021493.

2. Dugdale CM, Anahtar MN, Chiosi JJ, et al. Clinical, laboratory, and radiologic characteristics of patients with initial false-negative SARS-CoV-2 nucleic acid amplification test results. Open Forum Infectious Diseases 2020:ofaa559. https://doi.org10.1093/ofid/ofaa559.

3. Patrucco F, Carriero A, Falaschi Z, et al. COVID-19 diagnosis in case of two negative nasopharyngeal swabs: association between chest CT and bronchoalveolar lavage results. Radiology 2021:203776.

4. Fang Y, Zhang H, Xie J, et al. Sensitivity of Chest CT for COVID-19: Comparison to RT-PCR. Radiology 2020:200432. https://doi.org10.1148/radiol.2020200432.

5. Ai T, Yang Z, Hou H, et al. Correlation of Chest CT and RT-PCR Testing in Coronavirus Disease 2019 (COVID-19) in China: A Report of 1014 Cases. Radiology 2020:200642. https://doi.org10.1148/radiol.2020200642.

6. Schalekamp S, Bleeker-Rovers CP, Beenen LFM, et al. Chest CT in the Emergency Department for Diagnosis of COVID-19 Pneumonia: Dutch Experience. Radiology 2020:203465.

7. Som A, Lang M, Yeung T, et al. Implementation of the Radiological Society of North America Expert Consensus Guidelines on Reporting Chest CT Findings Related to COVID-19: A Multireader Performance Study. Radiology: Cardiothoracic Imaging 2020;2(5):e200276.

8. Simpson S, Kay FU, Abbara S, et al. Radiological Society of North America Expert Consensus Statement on Reporting Chest CT Findings Related to COVID-19. Endorsed by the Society of Thoracic Radiology, the American College of Radiology, and RSNA. Radiology: Cardiothoracic Imaging 2020;2(2):e200152.

9. Stephanie S, Shum T, Cleveland H, et al. Determinants of chest x-ray sensitivity for covid-19: A multi-institutional study in the united states. Radiology: Cardiothoracic Imaging 2020;2(5):e200337.

10. Little BP. False-Negative Nasopharyngeal Swabs and Positive Bronchoalveolar Lavage: Implications for Chest CT in Diagnosis of COVID-19 Pneumonia. Radiological Society of North America; 2021.

11. Dugdale CM, Rubins DM, Lee H, et al. Coronavirus Disease 2019 (COVID-19) Diagnostic Clinical Decision Support: A Pre-Post Implementation Study of CORAL (COvid Risk cALculator). Clinical Infectious Diseases 2021. https://doi.org10.1093/cid/ciab111.

12. Volpicelli G, Gargani L. Sonographic signs and patterns of COVID-19 pneumonia. The Ultrasound Journal 2020;12(1):1–3.

13. Huang Y, Wang S, Liu Y, et al. A preliminary study on the ultrasonic manifestations of peripulmonary lesions of non-critical novel coronavirus pneumonia (COVID-19). Available at SSRN 3544750 2020.

14. Smith MJ, Hayward SA, Innes SM, Miller ASC. Point-of-care lung ultrasound in patients with COVID-19–a narrative review. Anaesthesia 2020.

15. Peng Q-Y, Wang X-T, Zhang L-N, Chinese Critical Care Ultrasound Study G. Findings of lung ultrasonography of novel corona virus pneumonia during the 2019–2020 epidemic. Intensive care medicine 2020:1.

16. Soldati G, Smargiassi A, Inchingolo R, et al. Proposal for International Standardization of the Use of Lung Ultrasound for Patients With COVID-19: A Simple, Quantitative, Reproducible Method. Journal of Ultrasound in Medicine 2020.

17. Mento F, Perrone T, Macioce VN, et al. On the Impact of Different Lung Ultrasound Imaging Protocols in the Evaluation of Patients Affected by Coronavirus Disease 2019: How Many Acquisitions Are Needed? J Ultrasound Med. © 2020 American Institute of Ultrasound in Medicine.; 2020.

18. Pivetta E, Goffi A, Tizzani M, et al. Lung Ultrasonography for the Diagnosis of SARS-CoV-2 Pneumonia in the Emergency Department. Ann emerg med 2020.

19. Lieveld AWE, Kok B, Schuit FH, et al. Diagnosing COVID-19 pneumonia in a pandemic setting: Lung Ultrasound versus CT (LUVCT)–a multicentre, prospective, observational study. ERJ Open Research 2020;6(4).

20. Haak SL, Renken IJE, Jager LC, Lameijer H, van der Kolk BYM. Diagnostic accuracy of point-of-care lung ultrasound in COVID-19. Emergency Medicine Journal 2021;38(2):94. https://doi.org10.1136/emermed-2020-210125.

21. Lichtenstein DA. Lung ultrasound in the critically ill. Annals of Intensive Care 2014/01/09 2014;4(1):1. https://doi.org10.1186/2110-5820-4-1.

22. Simpson S, Kay FU, Abbara S, et al. Radiological society of north America expert consensus document on reporting chest CT findings related to COVID-19: endorsed by the society of thoracic Radiology, the American college of Radiology, and RSNA. Radiology: Cardiothoracic Imaging 2020;2(2):e200152.

23. DeLong ER, DeLong DM, Clarke-Pearson DL. Comparing the areas under two or more correlated receiver operating characteristic curves: a nonparametric approach. Biometrics Sep 1988;44(3):837–45.

24. McKinney W. Data structures for statistical computing in python. Austin, TX. p. 51–56.

