## Appendix for "A Simplified Point-of-Care Lung Ultrasound Protocol to Detect Coronavirus Disease 2019 in Inpatients: A Prospective Observational Study"

### Figure 1: POCUS Reader Scoring Form

| Select ONE per Zone, leave blank if none apply | R1 | R2 | R3 | R4 | L1 | L2 | L3 | L4 | R5 | R6 | L5 | L6 |
|---|---|---|---|---|---|---|---|---|---|---|---|---|
| TLS / cannot interpret | ☐ | ☐ | ☐ | ☐ | ☐ | ☐ | ☐ | ☐ | ☐ | ☐ | ☐ | ☐ |
| normal lung / A-lines | ☑ | ☑ | ☑ | ☐ | ☑ | ☑ | ☐ | ☑ | ☑ | ☑ | ☐ | ☑ |
| 1-2 B-lines | ☐ | ☐ | ☐ | ☐ | ☐ | ☐ | ☑ | ☐ | ☐ | ☐ | ☑ | ☐ |
| 3+ B-lines | ☐ | ☐ | ☐ | ☐ | ☐ | ☐ | ☐ | ☐ | ☐ | ☐ | ☐ | ☐ |
| Confluent B-lines ≤50% intercostal space | ☐ | ☐ | ☐ | ☑ | ☐ | ☐ | ☐ | ☐ | ☐ | ☐ | ☐ | ☐ |
| Confluent B-lines >50% intercostal space | ☐ | ☐ | ☐ | ☐ | ☐ | ☐ | ☐ | ☐ | ☐ | ☐ | ☐ | ☐ |
| **Pleural Line Irregularity (Select ONE per Zone)** | R1 | R2 | R3 | R4 | L1 | L2 | L3 | L4 | R5 | R6 | L5 | L6 |
| Normal/regular pleural line (or small indent only) | ☑ | ☑ | ☑ | ☑ | ☑ | ☑ | ☑ | ☑ | ☑ | ☑ | ☐ | ☑ |
| Broken / irreg pleural line in ≤50% intercostal space | ☐ | ☐ | ☐ | ☐ | ☐ | ☐ | ☐ | ☐ | ☐ | ☐ | ☑ | ☐ |
| Irregular pleural line covering >50% intercostal space | ☐ | ☐ | ☐ | ☐ | ☐ | ☐ | ☐ | ☐ | ☐ | ☐ | ☐ | ☐ |
| **Consolidation (Select ONE per Zone** | R1 | R2 | R3 | R4 | L1 | L2 | L3 | L4 | R5 | R6 | L5 | L6 |
| no consolidation | ☑ | ☑ | ☑ | ☐ | ☑ | ☑ | ☑ | ☑ | ☑ | ☐ | ☐ | ☑ |
| small (< 1cm) sub-pleural consolidation | ☐ | ☐ | ☐ | ☑ | ☐ | ☐ | ☐ | ☐ | ☐ | ☐ | ☑ | ☐ |
| larger (>1cm) subpl.consol./shred sign | ☐ | ☐ | ☐ | ☐ | ☐ | ☐ | ☐ | ☐ | ☐ | ☑ | ☐ | ☐ |
| hepatization with air bronchograms | ☐ | ☐ | ☐ | ☐ | ☐ | ☐ | ☐ | ☐ | ☐ | ☐ | ☐ | ☐ |

================================

**Is there a Right pleural effusion?**
* must provide value

- ○ no
- ○ trace/minimal
- ◉ small
- ○ moderate
- ○ large

reset

**Is there a Left pleural effusion?**
* must provide value

- ○ no
- ○ trace/minimal
- ○ small
- ○ moderate
- ◉ large

**Figure 2: Study Population**

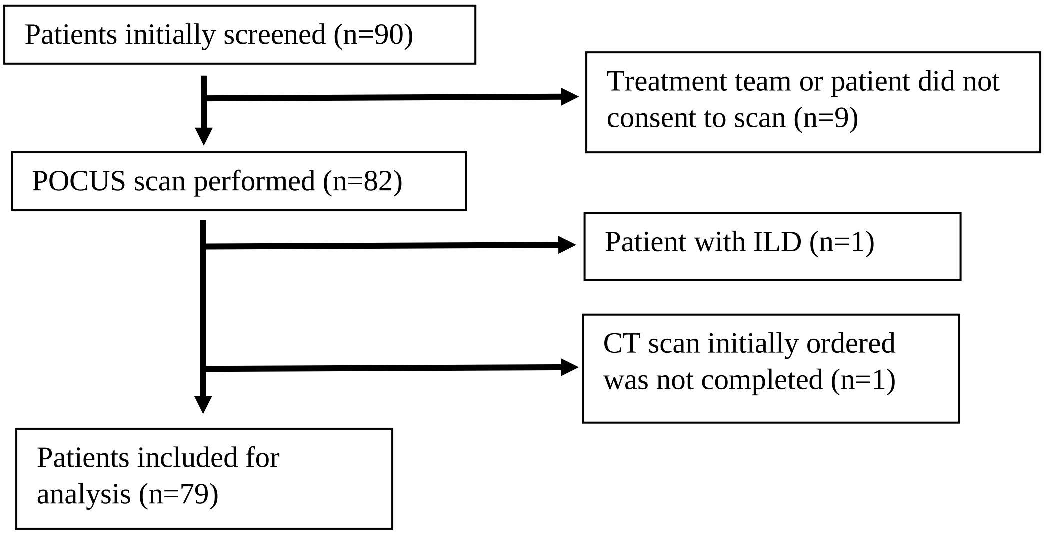

**Figure 3: CT, CXR, and POCUS Interpretations with NAT Results**

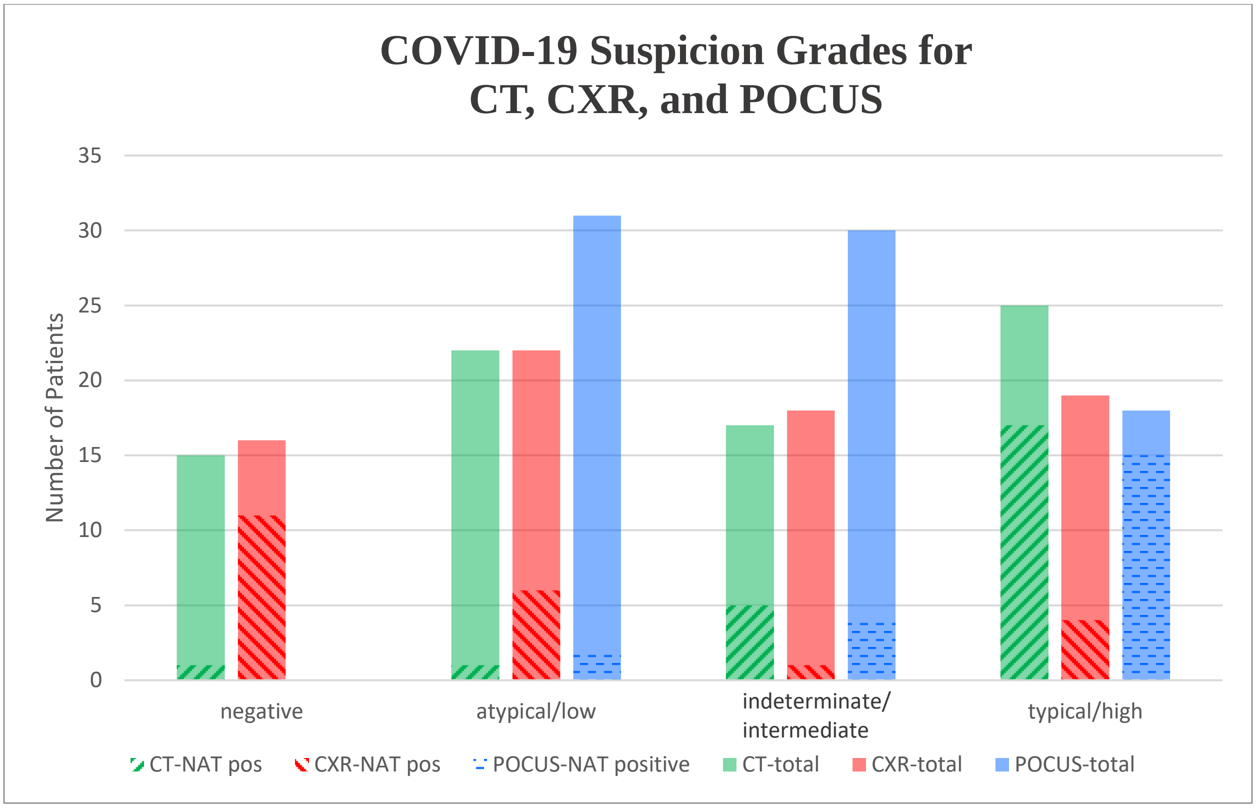


**Legend:** Patients categorized by imaging interpretation (e.g., green bars indicate how patients were categorized by CT interpretation, with diagonal marks indicating the patients who also tested positive for COVID-19 on NAT). POCUS here indicates the simplified four-zone protocol.

**Figure 4: Feature Importance of Individual Lung Findings for Typical CT Pattern, Using Lasso Regression**

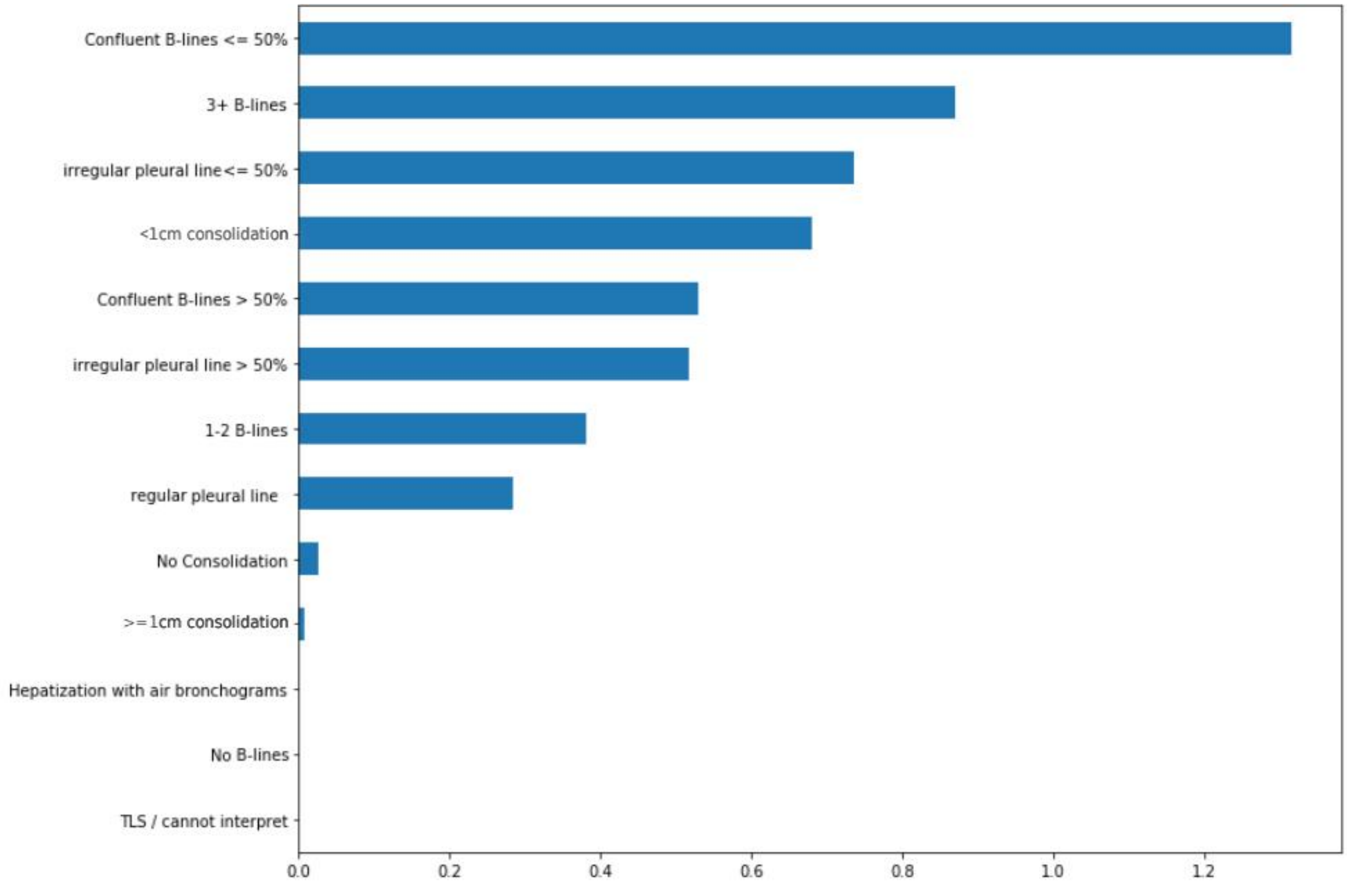

**Figure 5: Feature Importance of Individual Lung Zones for Typical CT Pattern, Using Random Forest**

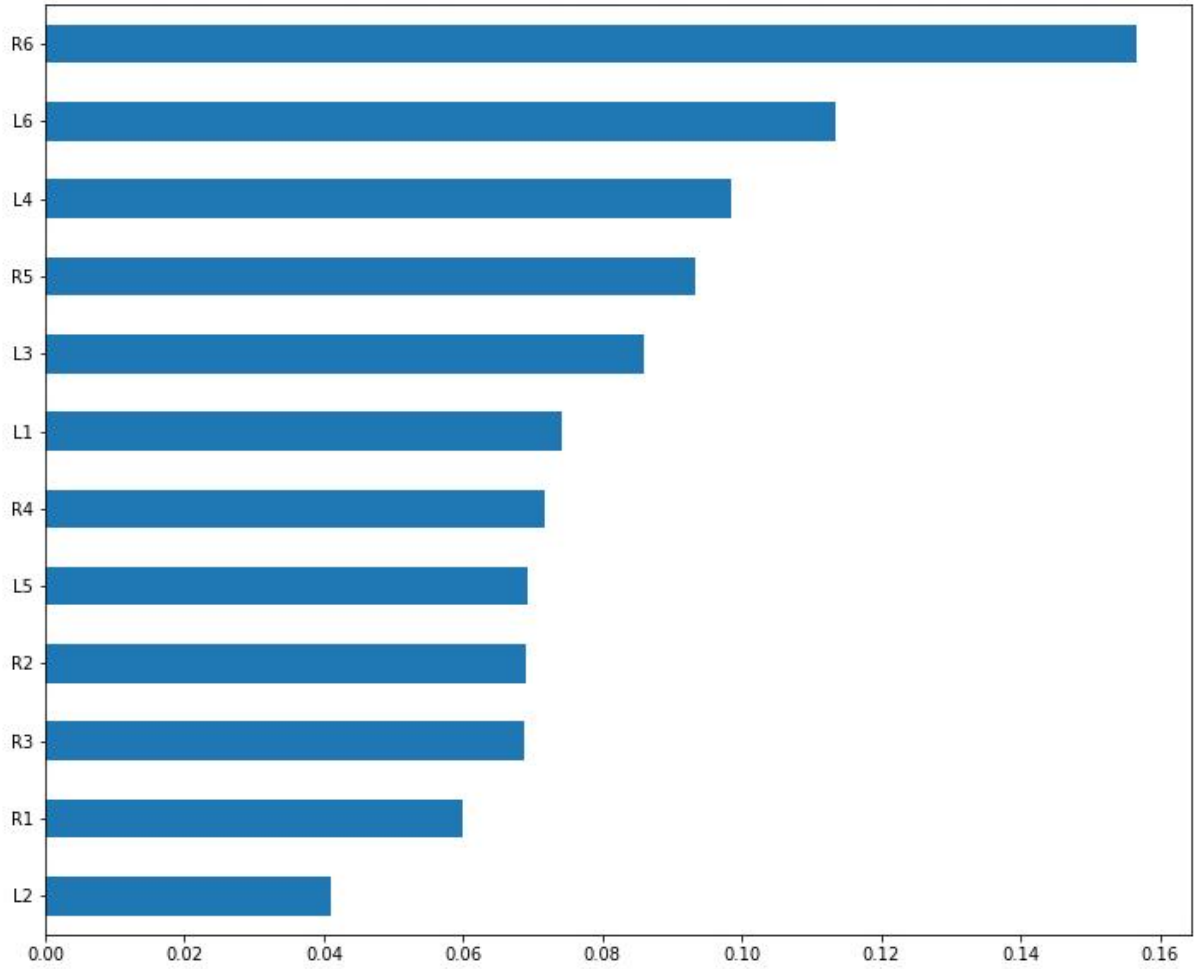
